# Racial Variation in Cerebral Near Infrared Spectroscopy Accuracy Among Infants in a Cardiac Intensive Care Unit

**DOI:** 10.1101/2025.04.21.25326146

**Authors:** Callie Marshall, Stephanie Diggs, Morgan Pfeiffer, Anna Gerst, Alexa Brumfiel, Zachary Vesoulis

## Abstract

**Objective:** Pulse oximeters overestimate arterial oxygen saturations in black versus white adults, children, and infants. While race’s impact on near-infrared spectroscopy (NIRS) accuracy is less studied, some adult research suggests decreased accuracy in black patients. This study investigates the effect of race on NIRS accuracy in infants in a cardiac intensive care unit (CICU).

**Study Design:** A retrospective chart review was conducted for infants admitted to St. Louis Children’s Hospital CICU from 2017-2023. Bland-Altman plots, Pearson correlations, and mean biases were analyzed.

**Result:** 254 infants (13% Black, 87% White) provided 3,687 central venous oxygen saturation (ScvO_2_)-cerebral regional oxygen saturation (rScO_2_) pairs. Measurement bias was −3.2% in Black infants and +0.1% in White infants (p<0.01).

**Conclusion:** Cerebral NIRS underestimates ScvO_2_ in Black infants but maintains minimal measurement bias in White infants. This is the first study to assess race and NIRS accuracy in infants; the difference is statistically significant but not clinically relevant in most contexts.

## Introduction

In the late 1970’s, Dr. Frans Jöbsis described the use of a near infrared spectroscopy (NIRS) monitor in a feline model to non-invasively measure oxygenated and deoxygenated hemoglobin in the brain, thus setting the framework for characterizing the regional oxygen saturation (rSO_2_) of a target organ.^1^ Since then, NIRS monitors have gained regulatory approval and are commercially available for adult, pediatric, and neonatal applications. The clinical use of NIRS has grown and become the standard of care in some settings and institutions, especially pediatric cardiac critical care.^2,3^ Previous studies have established a strong correlation between the venous-weighted oxygen saturation estimate provided by NIRS and invasive measurement of central venous oxygen saturation (ScvO_2_).^4,5^ This indicator of the delicate balance between oxygen supply and demand can provide a real-time assessment of inadequate oxygenation, an occurrence which may lead to catastrophic outcomes including irreversible end organ damage and death.^4^ Studies of cerebral NIRS in infants with congenital heart disease (CHD) suggest an association between cerebral hypoxia and outcomes ranging from short-term morbidity to longer-term neurodevelopment, emphasizing the clinical value of this neuromonitoring modality.^6,7^

NIRS and other optical measures of oxygen saturation rely on the difference in light absorption between oxyhemoglobin and deoxyhemoglobin, employing proprietary algorithms to convert the ratio of emitted to absorbed light at two or more wavelengths into an estimate of oxygen saturation.^8^ However, melanin also absorbs light in the red and near-infrared light wavelengths used in these technologies.^8,9^ This competitive light absorption is not accounted for in commercial algorithms, leading to concerns about the potential for race-based disparities in performance. Subsequent studies of pulse oximeters have found a consistent overestimation of true oxygen saturation in patients with darker skin,^10,11^ a trend also identified in pediatric and neonatal patients.^12,13^ While NIRS monitoring can be distinguished from pulse oximetry by the use of light reflectance rather than transmission, measurement of all vascular compartments, and the use of different wavelengths of light, there is sufficient overlap in underlying methods to raise concern about inaccuracy. Currently, there are no clinical studies exploring the impact of skin pigment and pediatric or infant NIRS, but a small number of reports in the adult literature suggests that race-based performance disparity does exist.^14–16^

Given the growing clinical role of NIRS in infants with CHD, identifying any racial bias is paramount in ensuring equitable care in this vulnerable population. In this retrospective study, we identified a cohort of infants under 1 year of age in a cardiac intensive care unit (CICU) who underwent routine cerebral NIRS monitoring and had simultaneous collection of central venous saturation samples measured via co-oximetry. We hypothesized that differences in melanin concentration between Black and White infants would lead to systematic error in the ability of cerebral NIRS values to approximate ScvO_2_.

## Methods

### Cohort development

All infants admitted to the St. Louis Children’s Hospital (SLCH) CICU from October 2017 – January 2023 in their first year of life (age <365 days) were eligible for inclusion. Infants undergo routine monitoring of both cerebral and renal NIRS in the SLCH CICU (INVOS 5100c, Medtronic, Boulder CO). A central server (T3, Etiometry, Boston MA) prospectively archives all physiologic data for clinical and research use. Infants were included in the study if they had an ScvO_2_ measurement recorded during a period where they were also undergoing cerebral NIRS monitoring. Participants were excluded if a) they were >365 days of age at the time of measurement, b) ScvO_2_ measurements were not taken during the admission, c) cerebral NIRS monitoring was not being performed at the time of the blood gas, or d) cerebral NIRS data was missing, or the recording was corrupted at the time of blood gas.

Clinical data was extracted via retrospective chart review and included age at admission, requirement for extracorporeal membranous oxygenation (ECMO), and length of hospital and CICU stay, and an anatomical description of the cardiac anomaly from echocardiography. Infants were classified as Black or White based on parental identification on birth certificates. To avoid statistical imbalance, infants of Hispanic ethnicity and/or other races were excluded from analysis as they represent a small proportion of admitted patients (<5%).

To compare the severity of cardiac pathologies, the Society of Thoracic Surgeons (STS) – European Association for Cardio-Thoracic Surgery Congenital Heart Surgery Mortality Categories (STAT Category) were extracted from the institutional database. These categories range from 1 (lowest mortality) to 5 (highest mortality) and describe the mortality risk of a patient’s cardiac physiology and surgical intervention.^17^ Patients who did not undergo surgery did not have a STAT mortality score and were categorized as non-operative. This study was reviewed and approved by the IRB at Washington University under waiver of consent and was performed in accordance with the Declaration of Helsinki.

### ScvO_2_ assessment

While NIRS validation studies tend to assume a fixed ratio of venous to arterial blood volume, obtaining simultaneous oxygen saturations from both sites,^18^ further work has noted a close correlation between cerebral rSO_2_ and venous oxygen saturations obtained from both the jugular bulb (SjvO_2_) and central venous sites (ScvO_2_).^4,5^

Infants in the CICU routinely require central venous access for lab draws and medication/intravenous fluid administration. Central venous oxyhemoglobin saturations were drawn from these central catheters using aseptic technique with a minimum collection volume of 0.3 milliliters (mL). Samples were placed in an air-tight balanced heparinized blood gas syringe and sent to the clinical laboratory within 5 minutes of collection. A gas analyzer (ABL800 Flex, Radiometer America, Brea, CA) measured oxyhemoglobin content to provide a central venous saturation (ScvO_2_). Sample collection times were contemporaneously entered into the electronic medical record (EMR) at the time of blood draw and this timing data was retrieved during chart review.

### rSO_2_ processing

Raw physiologic recording files were exported from the local T3 database in comma separated value (CSV) format; all data were sampled at 0.2 Hz (once every 5 seconds). A custom processing pipeline was developed in Python. In the first stage, it cleaned, aligned, de-identified, and consolidated multiple days of raw recordings into a single file per patient. The second phase of this pipeline used the electronic timestamp of ScvO_2_ sample acquisition recorded in the EMR to identify a time-matched 60-second window of cerebral rSO_2_ (rScO_2_) data centered on the ScvO_2_ sample acquisition time point. The entire 60-second window was then averaged to minimize the effect of transient variations and/or motion artifact contamination in rScO_2_ measurements. Thus, for each ScvO_2_ sample, this pipeline yielded a single data triplet (postnatal age in days, ScvO_2_, rScO_2_) for use in the remainder of the analysis.

### NIRS performance metrics

Performance standards for non-invasive tissue oximeters are primary set by ISO 80601-2-85, currently in the first edition, which has been adopted by FDA amongst other regulatory bodies. Similar to pulse oximeter validation, performance is assessed using several different metrics including simple scatter plot analysis, proportional bias (Bland-Altman), accuracy (measured via average root mean squared), and mean bias. This validation standard requires variance by no greater than 10% from a blood reference value across the range of typically experienced tissue oxygen saturation levels, from 50 to 85%. Validation is conducted during controlled desaturation testing against the reference standard of SavO_2_, where *SavO_2_ = R × SaO_2_ + (1 - R) × SjvO_2_*. This formula uses the arterial oxygen saturation (SaO_2_) and jugular venous oxygen saturation (SjvO_2_) along with a manufacturer-defined ratio (R) of venous to arterial blood (typically 70/30) to calculate mixed venous oxygen saturation.

In this study, we adopt a modification of same performance framework in the ISO standard. As repeated sampling of jugular venous saturation in neonates is technically challenging and poses excessive risk, a surrogate measure of mixed venous saturation (ScvO_2_ via co-oximetry) from a central venous catheter is used as an alternate reference standard.

Performance metrics evaluated included: Mean bias (B) – This method calculates the average difference between rSO_2_ and ScvO_2_ using the formula 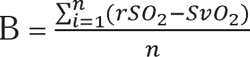. Positive values indicate an overestimate of central venous saturation by NIRS, while negative values denote underestimation.

Average root mean squared (A_rms_) – Capturing both accuracy and precision, A_rms_ is calculated as average difference between rScO_2_ and ScvO_2_ using the formula 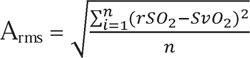. Arms, therefore, is always a positive value with a minimum value of 0 (no error) that increases as the degree of error increases.

Proportional bias – The Bland-Altman method plots differences between paired measurements (e.g., rScO_2_ and ScvO_2_) against their means, establishing a mean difference (bias) and 95% limits of agreement (mean ± 1.96 standard deviations). When proportional bias exists, the plot shows a significant slope rather than random scatter, indicating that one method increasingly over- or under-estimates compared to the other as measurement values increase.

### Statistical approach

Infant characteristics were compared using Fisher’s Exact Test for categorical variables and Mann-Whitney U test for continuous variables. The linear relationship between rSO_2_ and ScvO_2_ were modeled using the Pearson correlation coefficient and conventional linear regression. Smoothed conditional means were calculated via the ggplot2package for the R statistical package (R version 4.0.3, R Foundation for Statistical Computing, Vienna, Austria) utilizing the locally estimated scatterplot smoothing (LOESS) method.

## Results

### Study cohort description

A total of 687 infants were admitted to the CICU during the study period. Of these, 32 did not have a race of Black or White listed. Of the remaining infants, 216 did not have any ScvO_2_ values obtained during their admission while an additional 186 did not have a valid rSO_2_ during the period the ScvO_2_ value was obtained (Supplemental Figure 1). Ultimately, 3,687 rScO_2_-ScvO_2_ pairs were analyzed from 253 infants (87% White, 13% Black) (Table 1). The distribution of race by sample varied from underlying patient distribution, with 77% of samples coming from White patients and 23% of samples coming from Black patients. Although the two groups were similar in age at admission, Black infants had significantly lower mean rScO_2_ (58.6% v 68.2%, p<0.01) and ScvO_2_ (61.8% v 68.1%, p<0.01) values than White infants. Among Black infants, a statistically insignificant trend emerged towards longer length of stay in both the hospital (63 days v 40 days, p=0.05) and in the CICU (30 days v 17 days, p=0.09). STS STAT categories were similar between the two populations (p=0.84) as were the proportion of cyanotic cardiac lesions (p=0.72) and lesions with single ventricle physiology (p=0.59) (Table 2).

**Table 1.** Cohort descriptive statistics.

|  | Black Infants,<br>N=34 | White Infants, N=219 | p-value |
| --- | --- | --- | --- |
| NIRS Values, N (%) | 833 (23%) | 2854 (77%) |  |
| ECMO, N (%) | 6 (18%) | 24 (11%) | P=0.25 |
| Average rSO<sub>2</sub>, % (SD) | 58.6 (12.5) | 68.2 (12.1) | **P<0.01** |
| Average SvO<sub>2</sub>, % (SD) | 61.8% (13.3) | 68.1% (13.0) | **P<0.01** |
| Average Age at Admission, days | 87 | 88 | P=0.72 |
| Average Hospital Length of Stay, days | 63 | 40 | P=0.05 |
| Average CICU Length of Stay, days | 30 | 17 | P=0.09 |

**Table 2.** Cohort cardiac physiology descriptive statistics.

| Society of Thoracic Surgeons Surgical STAT Category | Black | White | P-value |
| --- | --- | --- | --- |
| 1 | 12 (35%) | 73 (33%) |  |
| 2 | 5 (15%) | 46 (21%) |  |
| 3 | 4 (12%) | 40 (18%) |  |
| 4 | 5 (15%) | 24 (11%) |  |
| 5 | 2 (6%) | 15 (7%) | P=0.84 |
| Non-operative | 6 (18%) | 21 (10%) | P=0.22 |
| Descriptive Cardiac Pathology |  |  |  |
| Ventricular Septal Defect/Atrial Septal Defect | 6 (18%) | 32 (15%) |  |
| Tetralogy of Fallot | 5 (15%) | 34 (16%) |  |
| Complete AV Canal | 3 (9%) | 21 (10%) |  |
| Transposition of the Great Arteries | 2 (6%) | 21 (10%) |  |
| Pulmonary Atresia | 2 (6%) | 5 (2%) |  |
| Hypoplastic Left Heart Syndrome | 2 (6%) | 11 (5%) |  |
| Aortic Arch Interruption/Coarctation | 2 (6%) | 26 (12%) |  |
| Cardiomyopathy | 2 (6%) | 5 (2%) |  |
| Tricuspid Atresia | 1 (3%) | 4 (2%) |  |
| Double Outlet Right/Double Inlet Left Ventricle | 0 | 15 (7%) |  |
| Total Anomalous Pulmonary Venous Connection | 0 | 2 (1%) |  |
| Truncus Arteriosus | 0 | 4 (2%) |  |
| Other | 9 (26%) | 39 (19%) |  |
| Single Ventricle Physiology | 3 (9%) | 30 (14%) | P=0.59 |
| Cyanotic | 14 (41%) | 98 (45%) | P=0.72 |

### Group NIRS Accuracy

rSO_2_ and ScvO_2_ measurements correlated moderately with a correlation coefficient of r=0.52 (Figure 1). The mean bias was calculated (rScO_2_ – ScvO_2_) such that a positive value indicates overestimation while negative shows underestimation. The mean bias of all samples was −0.6%, indicating a slight underestimation of the true central venous oxygen saturation. The 95% limits of agreement for the total population were −28.9% to 22.6% (Figure 1).

**Figure 1.**
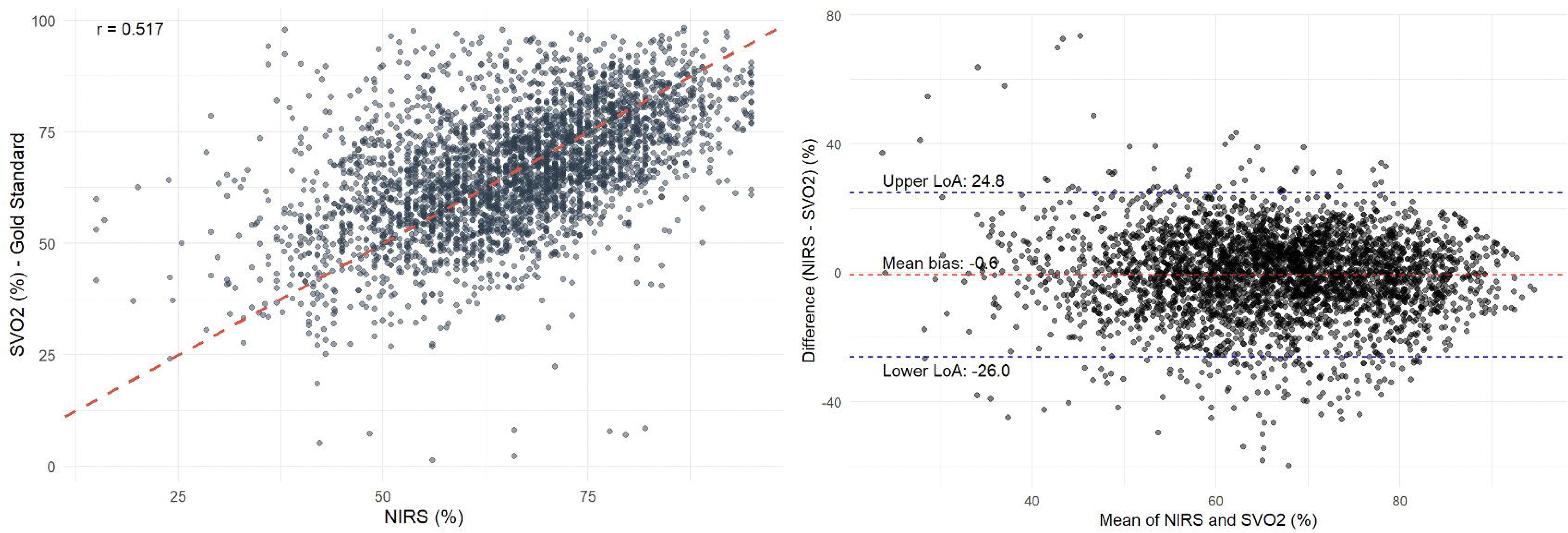
(Left) Pearson correlation plot comparing cerebral rSO_2_ (x-axis) and ScvO_2_ (y-axis). (Right) Bland-Altman plot for the cohort with mean bias and limits of agreement.

### Differences by Race

The mean bias was −3.2% for Black infants versus +0.1% in White infants (p<0.01), demonstrating an underestimation by cerebral NIRS of central venous oxygen saturation in Black infants and near perfect accuracy in White infants (Figure 2). A_rms_ was 13.5% in Black infants and 12.8% in White infants, also denoting increased error among Black patients. Bland-Altman analysis revealed upper and lower limits of agreement for Black patients (22.6 and −28.9 respectively) and White patients (25.2 and −25.0 respectively).

**Figure 2.**
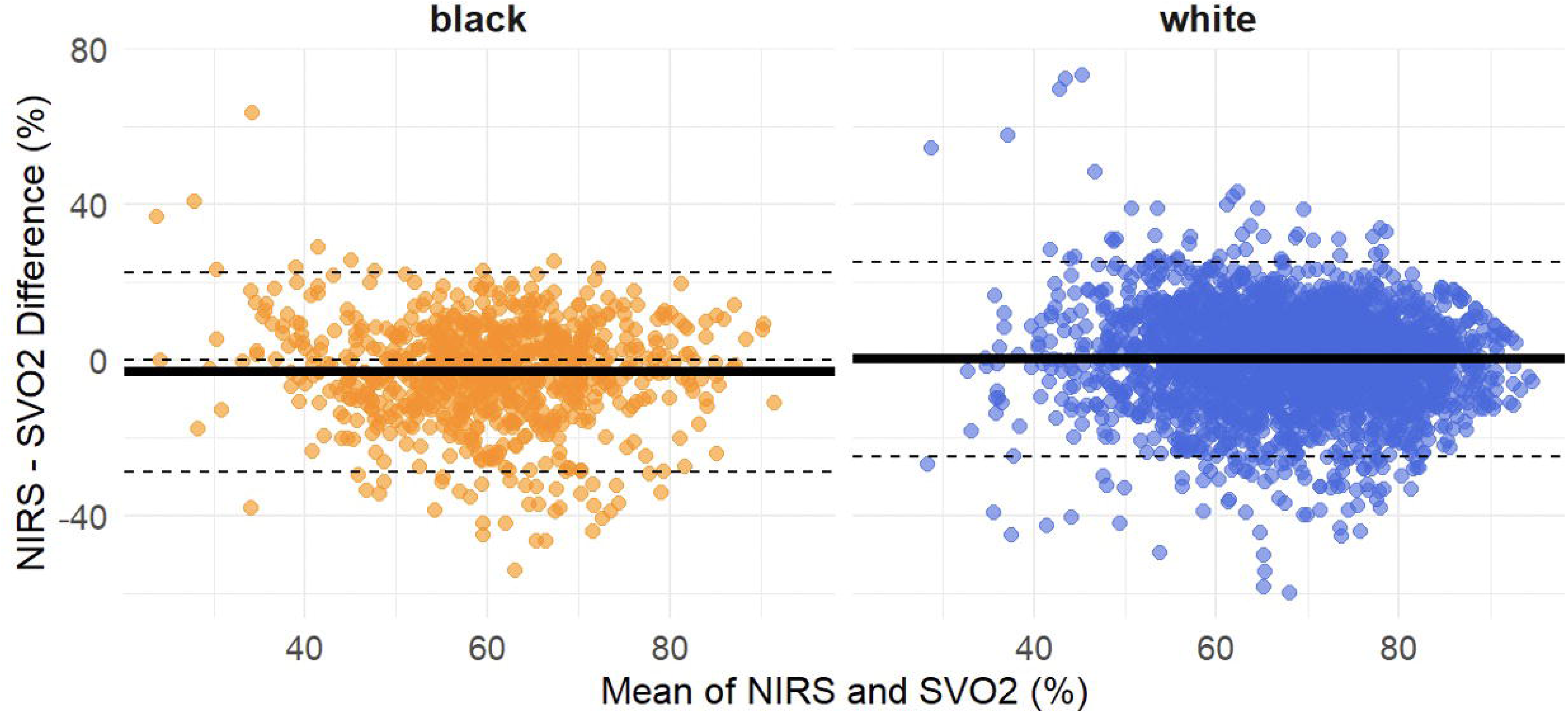
Bland-Altman plot for Black (left) and White (right) infants with mean bias (solid line) and limits of agreement (dotted lines).

### Age-based Reference Curve

Mean bias increased with increasing postnatal age from −1.8% (indicating underestimation of rScO_2_ values) soon after birth to +2.8% (denoting overestimation) in infants greater than 6 months of age. Despite the variation in cardiac anatomy and physiology amongst the cohort, the rScO_2_ and ScvO_2_ remained in the 60-70% range (Figure 3) for most time points while the average for specific age groups (0-30 days, 31-90 days, 91-180 days, 181-365 days) did not dip below 62% for either rSO_2_ or ScvO_2_ (Supplemental Table 1).

**Figure 3.**
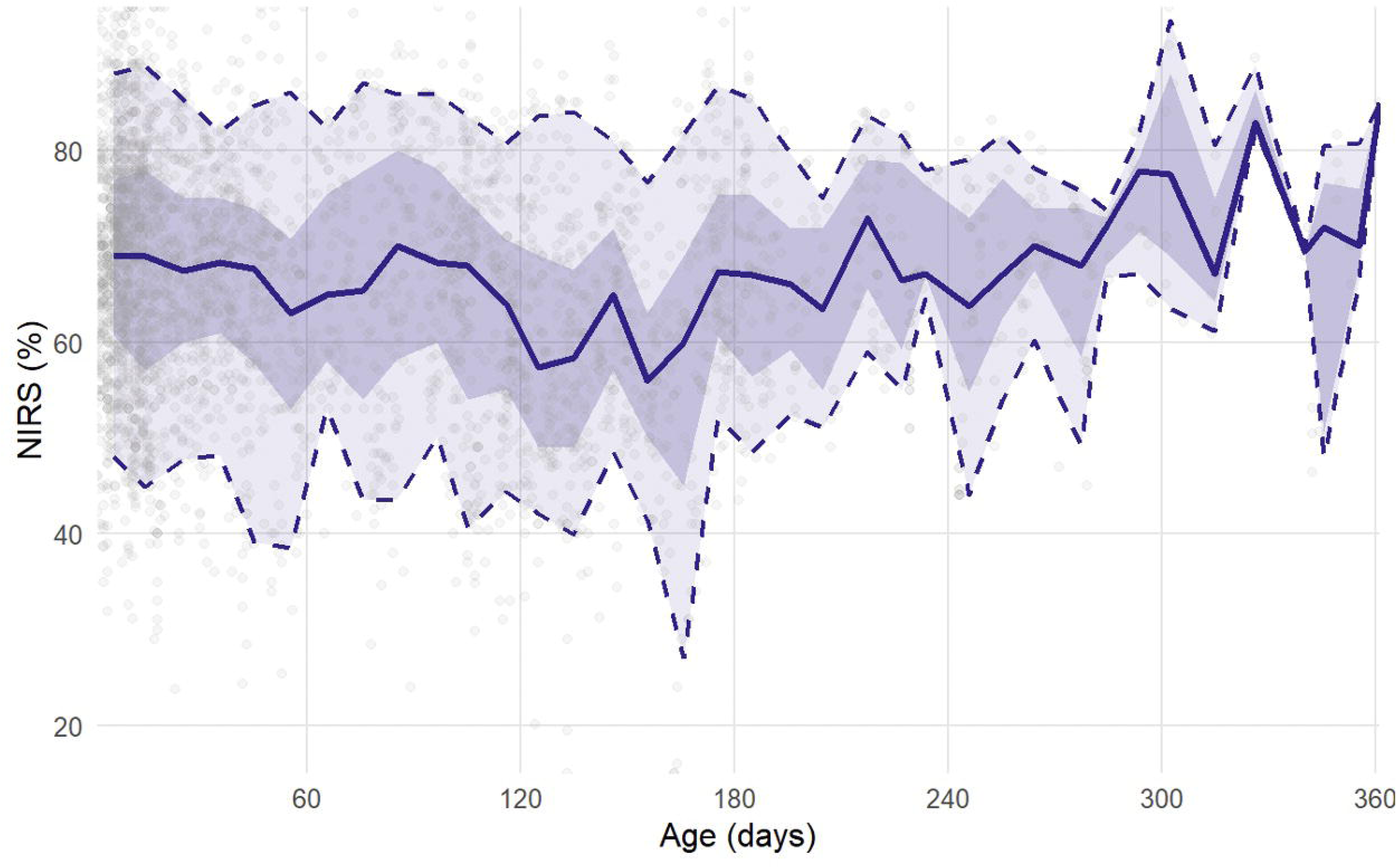
Locally-estimated scatterplot smoothing (LOESS) of rScO_2_ by postnatal age. Solid line represents median, shaded region encompasses 25^th^ to 75^th^ percentile while dotted line includes 5^th^ to 95^th^ percentile.

## Discussion

Cerebral NIRS modestly underestimates central venous oxygen saturation in Black infants in a cardiac intensive care unit while minimally overestimating central venous oxygen saturation among White infants. This study is the first to assess the impact of race on the accuracy of NIRS in the first year of life. Overall, the correlation between rScO_2_ and ScvO_2_ measurements was moderately positive. Finally, an age-based reference curve for CHD infants in the first year of life demonstrates a consistent range of cerebral saturations despite a diversity of cardiac anatomy and pre/post repair states.

Standards for brain-specific monitoring in infants with CHD vary by institution^6^ with 64% to 72% of European centers reporting routine NIRS monitoring in the pre- and post-operative periods, respectively.^3^ While a growing body of data supports an association between reduced cerebral rScO_2_ and poor neurodevelopmental outcomes^7^ and adverse events,^6^ cerebral rScO_2_ values considered normal vary by NIRS monitor and population. Healthy neonatal and infant cerebral rSO_2_ values average around 77% +/- 9% and 64% +/- 5% respectively^19,20^ while children with CHD had a similar average of 69% +/- 6%.^21^ Cerebral hypoxia thresholds for intervention have been difficult to establish in humans, but data from neonatal-equivalent animal models suggest a markedly increased incidence of brain injury when cerebral hypoxia events exceed 30 minutes below a saturation of 30-45%.^22,23^ Several studies suggested the use of a somewhat higher threshold of 50% to avoid adverse outcomes; a buffer between low-normal and critical cerebral saturations which offers clinicians a chance investigate and attempt to correct cerebral hypoxemia.^24^ NIRS-based intervention algorithms and goal-directed care strategies are in nascent stages in pediatric cardiac critical care, but one study observed a 27% reduction in the relative risk of mortality after providing a bedside “thought aid” to help guide medical decision making in the setting of decreased rScO_2_.^25^ At this stage, it is not clear if the mean bias difference we identified in Black versus White infants will affect current practice, but as research continues to explore goal-directed care and narrower intervention targets, this discrepancy may pose bias challenges.

Understanding the physiologic rationale for this racial discrepancy begins by appreciating that NIRS leverages differences in spectral absorption properties between oxyhemoglobin and deoxyhemoglobin in the 700-900 nm range.^8^ Light in this spectrum easily penetrates skin and bone to reach the underlying cerebral tissue.^26^ According to the Beer-Lambert law, both the concentrations and spectral absorption properties of chemical compounds in a measured solution determine the magnitude and wavelengths of absorbed light, properties which can be used to approximate the relative contribution of different chromophores to the overall absorption of light in a substance.^27^ The Beer-Lambert law has been modified and updated for tissue diagnostics, to account for challenges encountered in the clinical setting like light scattering by blood and for differential tissue reflectance.^27^

Melanin is the term given to a group of related molecules who have a principal purpose of photoprotection of the skin and eyes. Eumelanin is the pigment responsible for ultraviolet (UV)-absorption, while also acting as an antioxidant and free radical scavenger. While absorption of UV light is the primary biologic activity of eumelanin, it absorbs light across a broad spectrum, with greater absorption at red-infrared wavelengths (650nm to 1mm) than in the UV band.^8^ Melanin therefore serves as a competitive chromophore to hemoglobin, changing the magnitude of light returned to the sensor such that higher epidermal melanin concentration decreases the signal to noise ratio (SNR) and may decrease rScO_2_ values.^9,28^ Although NIRS sensors employ two source-detector distances, aiming to use the measurement at the shorter separation (and consequent smaller penetration depth) to correct for the impact of superficial (i.e., epidermal) layers on the return signal, pigmentation continues to exert an estimated effect of 2-15% on rScO_2_ values in phantom models of neonates, children and adults.^28^ Therefore, higher epidermal melanin concentration in Black infants appears to disrupt the anticipated relationship between returned light intensity and the calculated rScO_2_, leading to decreased accuracy in this population.

Although the pediatric and neonatal literature has not yet explored the impact of race on the accuracy of NIRS, studies in adults have identified a pattern also noted in our data: NIRS tends to either underestimate central or mixed venous saturation in Black patients, although these findings are inconsistent. A 2024 systematic review on the topic identified two studies for inclusion.^30^ Stannard and colleagues did not identify an independent effect of self-identification for any of 7 racial groups on pre-bypass rSO_2_ prior to elective surgery measured by a ForeSight senor.^31^ Sun et al, on the other hand, found that with the INVOS 5100c in adults undergoing predominantly emergency surgery, pre-induction rScO_2_ values were significantly lower in African American versus Caucasian patients after multilinear regression.^15^ Furthermore, a study of five different NIRS sensors found that in patients with darker skin, the probes tended to produce more negative bias (underestimation) in all sensors, but this was only statistically significant for the ForeSight sensor.^14^ It is worth noting that starting in 2021 the ForeSight monitor incorporated a 5^th^ wavelength into the probe design (685, 730, 770, 805, and 870 nm), at least in part to compensate for the competitive effect of melanin, and may have influenced the lack of difference appreciated in the Stannard et al study. Other studies of healthy volunteers in a controlled environment demonstrate either no bias^32^ or lower rSO_2_ among those with darker skin.^16,33,34^

While several major studies in pulse oximetry have noted a decrease in accuracy at lower saturations^32,35^ and increasing measurement bias by race at these lower levels,^10,11^ few published studies have commented on the presence of proportional bias in rSO_2_ measurement. Bickler et al noted that all 5 sensors tested had more positive mean bias during hypoxia and a phantom model found that one of two sensors tested demonstrated decreased accuracy at decreased rSO_2_ levels.^14,28^ However, at least one of the major studies in NIRS accuracy and race in adults did not identify issues with proportional bias, with a normal distribution of rSO_2_ error between races across the observed rSO_2_ range.^31^ In our study, Black infants had significantly lower average ScvO_2_ and rScO_2_ values than White infants. However, the Bland-Altman analyses by race in our study demonstrate a symmetric distribution of data points around the horizontal lines of mean bias. Accounting for the lack of proportional bias, the racial discrepancy in mean bias in our study is likely not the result of differences in ScvO_2_ between the groups.

The trend towards underestimation by NIRS departs significantly from the infant, pediatric, and even adult literature on pulse oximetry that suggests disproportionate *overestimation* of arterial oxygen saturation in Black versus White patients.^13,36^ The cause of this discrepancy in directionality is unclear. Some studies posit that the “spectral signature” of melanin more closely resembled deoxyhemoglobin, with a gradual smooth decline in absorption as wavelength increases, compared to the sudden drop in absorption beyond 600nm exhibited by oxyhemoglobin.^8^ his may cause the device to interpret a greater deoxyhemoglobin in patients with darker skin.^28^ However, given that the similarity in absorption profiles between these two chromophores also exists well into the red light range typically used in pulse oximetry, this theory does not fully explain the performance discrepancy in over-versus underestimation by two optical oximeters which share similar methodological underpinnings.

An evaluation of NIRS performance in infants must also consider the unique hemoglobin subtype, fetal hemoglobin (Hb F), present in this population. The composition of adult hemoglobin is typically about 97% Hb A with 1% Hb F.^37^ A term infant, on the other hand, has approximately 80% Hb F at birth, a concentration that falls steadily over the first several months of life such that by 6 months, Hb F accounts for only 5% with Hb A comprising the majority of the remaining hemoglobin.^38^ The absorption spectra of HbA and HbF are not identical in the near-infrared range.^39^ In our study, given the change in mean bias from negative −1.83% in neonates to +2.8% in babies older than 6 months, the shift in HbF must be considered as a confounder. However, a study of term and preterm infants found positive correlations between cerebral rSO_2_ and HbF concentration only in preterm infants.^40^ Furthermore, a 1991 study of 6 infants using both adult and fetal absorption coefficient profiles did not find a significant increase in error between the two,^39^ while later studies found no relationship between HbF concentration and NIRS-derived measurements in preterm infants like cerebral fractional oxygen excretion^41^ and cerebral tissue oxygenation index.^42^ Despite the biologic plausibility of decreasing HbF concentrations over time impacting NIRS accuracy, the limited body of literature for term infants has not identified a significant effect. Furthermore, the age at admission was similar between Black and White infants suggesting an equal distribution of postnatal ages unlikely to differentially impact HbF concentrations.

As noted earlier, the ISO validation standard for NIRS oximeters involves simultaneous venous samples from the jugular bulb^43^ in conjunction with arterial samples to calculate a mixed venous saturation. While this source has the greatest proximity to cerebral venous supply, obtaining samples from a central venous source like the superior or inferior vena cava or the cavoatrial junction is safer and more clinically feasible in the infant given the frequency of central line use in critically ill patients. Pediatric studies have identified a moderate correlation between cerebral rSO_2_ and central venous oxygenation with correlation estimates ranging from 0.58 to 0.7 in studies predominantly obtained from children and infants with CHD.^4,5^ A systematic review of NIRS validation including central venous and jugular sites in children with CHD found limits of agreement in bias ranging from −25.6 to 38.6.^4^ Our study found similarly large limits of agreement.

Infants in our study were classified as Black or White based on self-report in birth certificate documentation extracted from electronic medical records. This binary system of self-report fails to capture the spectrum of skin pigmentation. To better elucidate the effect of melanin, some studies of racial discrepancies in monitoring technologies employ more nuanced clinical evaluation systems such as the Fitzpatrick scale^16^ or Munsell color chart.^44^ Other studies, especially those with larger populations, included multiple racial identities^31^ or subjective pigmentation classifications of light, medium, or dark.^45^ To further minimize subjectivity, objective assessments of skin pigmentation include melanin indices^28^ obtained via reflectance measurements of different spectral bands or melanosome volume fraction,^29^ an established estimation of the quantity of melanosomes in a given skin thickness for use in phantom models.^46^

Our study has several limitations. First, the heterogeneity of sample sites contributes to greater bias in the overall sample. Although most central lines terminated in the cavoatrial junction, lower extremity peripherally inserted catheters may terminate in the caudal aspect of the inferior vena cava (IVC), thus sampling blood returning from the lower extremities (one study found that even IVC saturations correlated with cerebral NIRS, albeit weakly).^4^ Nevertheless, the primary goal of this study was to evaluate the difference in accuracy by race, thus this limitation is unlikely to preferentially affect Black or White infants.

Secondly, the retrospective, binary, and self-reporting nature of race classification prevents more rigorous, nuanced assessment of skin pigmentation. Birth certificates typically list the race of the mother,^47^ thus creating ambiguity in babies with parents of different races, but literature suggests that for White and Black infants, the birth certificate is highly sensitive for parent-reported race.^48^ Nevertheless, more quantitative direct measurement techniques are available such that prospective studies can measure melanin content to derive a more granular relationship between device performance and skin pigmentation outside of the complicated social constructs of race.

Finally, the number of Black infants in this study was significantly smaller than that of White infants, while the number of rScO_2_-ScvO_2_ values was proportionally greater for Black infants, indicating more repeated measurements. Although statistically the greater number of repeated measurements in Black infants should improve precision, the significant inherent intra-patient variability in NIRS measurements may have counterintuitively contributed to the decreased accuracy appreciated in this population. While the proportion of the of the study population that was Black (13.4%) is nearly identical to the US population estimate in 2016 (13.3%), there is an inherent class imbalance concern in this study.^49^ Future studies with enriched cohorts, especially with greater racial and ethnic diversity, will be important to identify nuances in device performance. Ultimately, however, quantitative measurement of pigmentation should serve as the primary covariate and not race.

In conclusion, we identified a statistically significant racial disparity in NIRS oximeter performance with underestimation of cerebral saturation compared to a central venous oxygen gold standard for infants admitted to the CICU in the first year of life. This is the first study to evaluate the impact of race on NIRS accuracy in infants or children and adds to the growing body of literature identifying racial disparities in light-based physiologic monitors, potentially compounding existing racial disparities in mortality and morbidity in infants with CHD.^50^ Although the degree of underestimation demonstrated in this study is not clinically significant under current clinical practice guidelines, this discrepancy may pose challenges as providers incorporate more nuanced NIRS thresholds in goal-directed care for this vulnerable population or NIRS becomes incorporated into AI-based predictive models.

## Author Contributions

CM collected the data and wrote the manuscript; AG, MP & AB helped with data collection; SD gave feedback on study design; ZV conceptualized the study, analyzed the data and edited the manuscript

## Data Availability

All data produced in the present study are available upon reasonable request to the authors

## Acknowledgements

Matthew Canter, Society of Thoracic Surgery database manager, for providing descriptive data of cardiac pathophysiology

