## Supplemental Table 1 for "Racial Variation in Cerebral Near Infrared Spectroscopy Accuracy Among Infants in a Cardiac Intensive Care Unit"

Supplemental Table 1. rSO_2_ and ScvO_2_ by age group

| Age Group | N | rSO2 Mean (SD) | SvO2 Mean (SD) | Mean Bias (%) | 95% CI |
| --- | --- | --- | --- | --- | --- |
| 0-30 days | 1456 | 67.9 (12.7) | 69.7 (13.3) | -1.83 | (-2.46, -1.2) |
| 31-90 days | 700 | 65.9 (12.9) | 67.0 (12.6) | -1.17 | (-2.08, -0.26) |
| 91-180 days | 1019 | 62.6 (13.3) | 62.4 (13.5) | 0.15 | (-0.72, 1.02) |
| 181-365 days | 385 | 67.9 (10.3) | 65.1 (12.0) | 2.83 | (1.61, 4.06) |
