## Supplementary figures and images for "Racial Variation in Cerebral Near Infrared Spectroscopy Accuracy Among Infants in a Cardiac Intensive Care Unit"

### Supplemental Figure 1

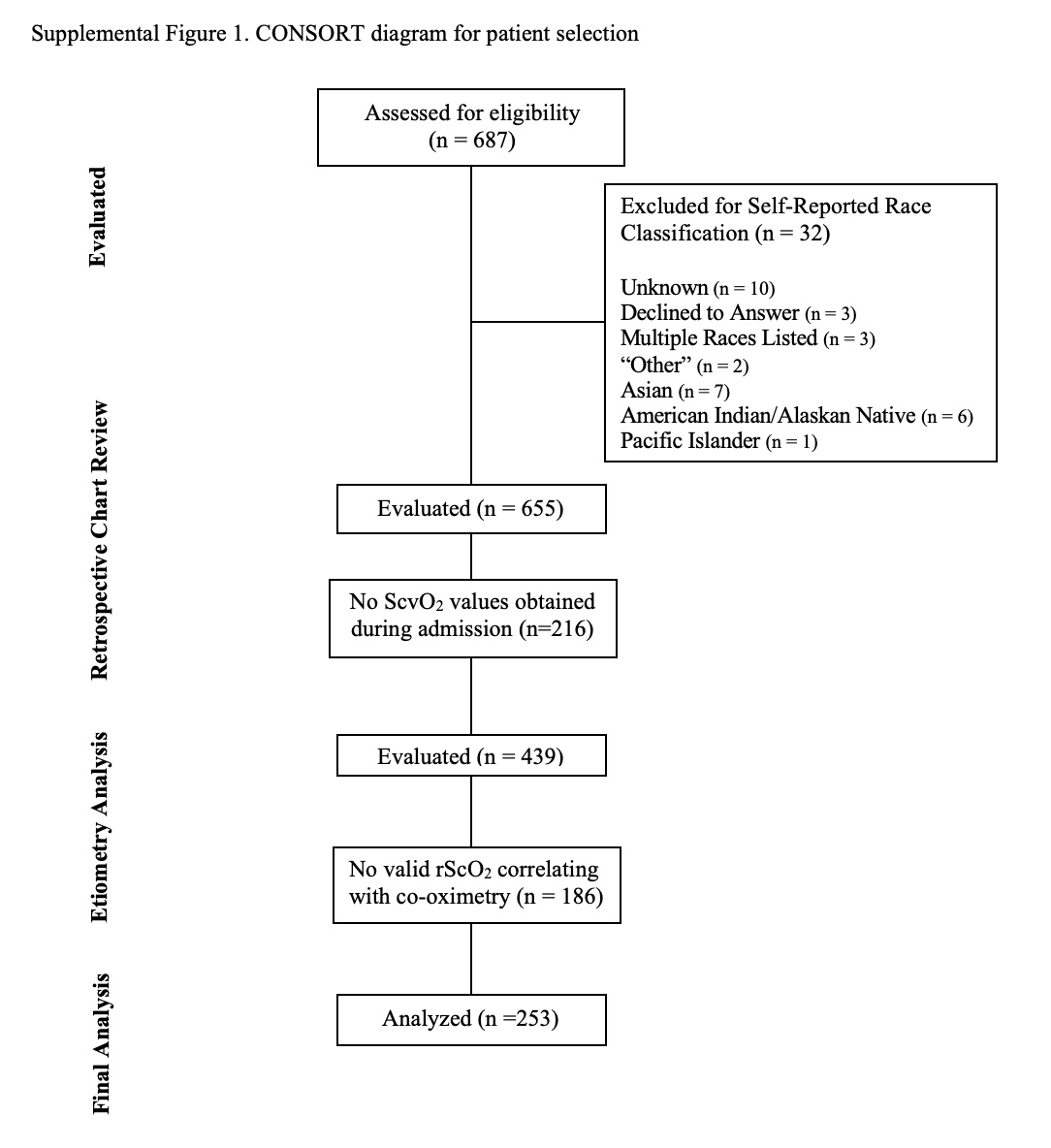
